# Large Language Model-Driven Evaluation of Medical Records Using MedCheckLLM

**DOI:** 10.1101/2023.11.01.23297684

**Authors:** Marc Cicero Schubert, Wolfgang Wick, Varun Venkataramani

## Abstract

Large Language Models (LLMs) offer potential in healthcare, especially in the evaluation of medical documents. This research introduces MedCheckLLM, a multi-step framework designed for the systematic assessment of medical records against established evidence-based guidelines, a process termed ‘guideline-in-the-loop’. By keeping the guidelines separate from the LLM’s training data, this approach emphasizes validity, flexibility, and interpretability. Suggested evidence-based guidelines are externally accessed and fed back into the LLM for a evaluation. The method enables implementation of guideline updates and personalized protocols for specific patient groups without retraining. We applied MedCheckLLM to expert-validated simulated medical reports, focusing on headache diagnoses following International Headache Society guidelines. Findings revealed MedCheckLLM correctly extracted diagnoses, suggested appropriate guidelines, and accurately evaluated 87% of checklist items, with its evaluations aligning significantly with expert opinions. The system not only enhances healthcare quality assurance but also introduces a transparent and efficient means of applying LLMs in clinical settings. Future considerations must address privacy and ethical concerns in actual clinical scenarios.

## Large Language Model-Driven Evaluation of Medical Records Using MedCheckLLM

Large Language Models (LLMs) have been utilized across a multitude of applications, demonstrating enormous potential in processing and comprehending complex datasets in healthcare^1^. One area yet to be thoroughly explored is the application of LLMs for the reliable and reproducible evaluation of medical documents. Automatic evaluation of these documents, if achieved effectively, has the potential to improve healthcare, enhance patient safety, reduce the risk of cognitive and other biases, and refine the training process of LLMs. Importantly, it is essential that the system’s reasoning process is a) transparent and comprehensible to human evaluators such as a checklist completion, and b) is guided by established medical guidelines proven to increase patient safety^2^ and the gold standard for implementing clinical care, thereby elevating the overall performance and applicability of AI-driven healthcare.

In this study, we introduce a framework which is based on a multi-step approach for medical record evaluation that incorporates guidelines directly into the evaluation process, a concept we term ‘guideline-in-the-loop’. Our proposed algorithm, named MedCheckLLM, is an LLM-driven structured, layered reasoning mechanism (Figure 1) designed to automate the evaluation of medical records, with a particular emphasis on the evaluation against evidence-based guidelines. Crucially, the guidelines are deterministically accessed by the LLM as out-of-training data. This rigorous separation of LLM and guidelines is expected to lead to increased validity and interpretability of the evaluations and offers flexibility for updating guidelines. The primary objective of this research is to introduce the conceptual framework and assess its feasibility. This approach is expected to have significant implications on healthcare quality and the transparent and efficient application of LLMs in clinical settings.

**Figure 1.**
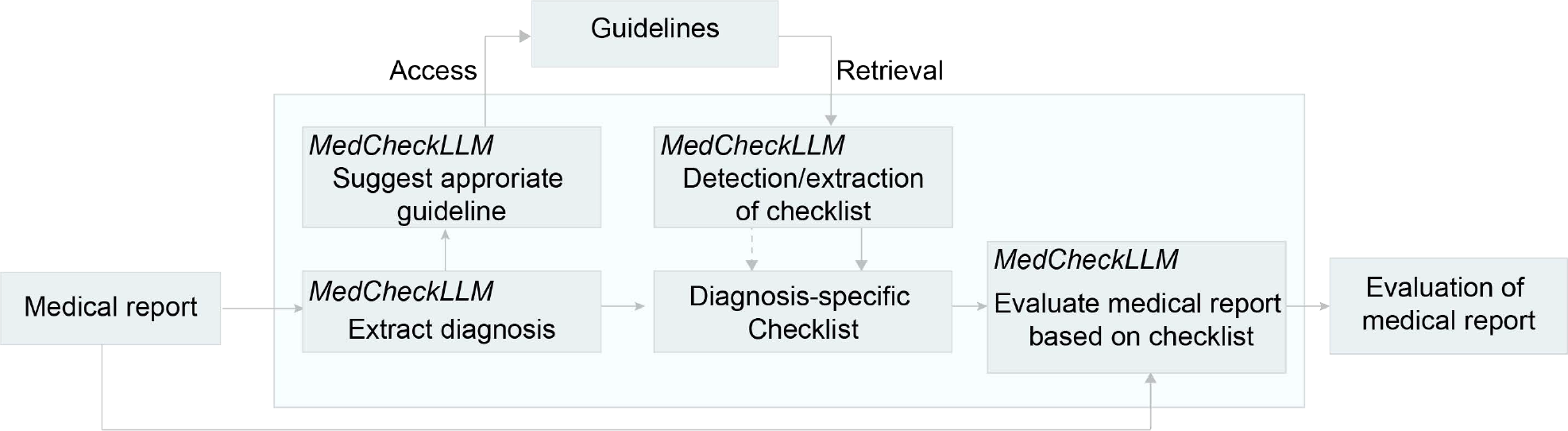
Workflow of MedCheckLLM.

## Methods

In this study, we utilized expert-validated simulated medical reports, which included patients’ medical history, examination outcomes, diagnosis, and treatment strategies. The MedCheckLLM algorithm begins by extracting a patient’s diagnosis from the medical report. Based on the extracted diagnosis, a suggestion for a guideline from appropriate medical societies is made. Guidelines are then accessed independently from the LLM’s mechanisms. Subsequently, guidelines are provided as input to the LLM and either identified as already formatted in a usable checklist or are converted into a checklist. Finally, this diagnosis-specific checklist is employed to assess the medical report. In this study, we evaluated whether the system is able to evaluate patient histories with headaches as leading symptom based on the structured guidelines from the International Headache Society. These checklists serve as benchmarks, used to assess the completeness, compliance, and possible inaccuracies of the diagnosis. The validity of this method was further confirmed by an evaluation of correct and incorrect synthetic doctor’s notes. The LLM “gpt-4-0613” was accessed using the Open-AI API. The study was conducted between July 24^th^ and August 30^th^ 2023.

## Results

We evaluated the medical report analysis conducted by MedCheckLLM for the diagnosis of various headache cases. MedCheckLLM extracted the specified diagnosis correctly in 100 % of cases, choosing from a list of 61 possible diagnoses from the ICHD-3^3^ (n=17 medical reports). The model suggested appropriate and existing evidence-based guidelines in 70.59 % (n=12 of 17) of medical reports. It could convert the format of the guidelines to checklists in 100 % of cases (n=17). MedCheckLLM accurately evaluated 87.0 % (67 of 77) of the checklist items. The explanations provided by MedCheckLLM for its evaluations were mostly in high agreement with expert evaluations, affirming its effectiveness. MedCheckLLM was able to identify incorrect diagnoses where the stated condition did not align with the rest of the content in 94.1% (16 of 17) doctor’s letters, while it correctly recognized 100% of the correct letters (n=17).

## Discussion

The framework of MedCheckLLM represents a promising approach for comprehensive, guideline-anchored review of electronic health records.

It holds the potential to advance healthcare by acting as a quality assurance tool, thereby improving individual provider assessment and patient-specific care due to its several advantages of distinct partitioning of the LLM and the guidelines over training guidelines into an LLM: The flexibility of the approach allows for immediate implementation of guideline updates as well as the option to implement specific and customized protocols for subgroups of patients, without the necessity of retraining. The interpretability makes it easy to understand why the algorithm made a particular decision, which is crucial in healthcare settings where explainability is required^4^. Additionally, they do not show bias based on the data that is presented to the LLM, reducing the risk of overfitting. Further, this approach can be used to evaluate the quality of electronic healthcare records that were previously used to train LLMs without extensive quality controls^5^. Well-structured guidelines are key to leveraging this process effectively. The framework facilitates improved data mining practices in electronic health records^5^, promotes healthcare quality assurance, and can contribute to a relevant leap forward in healthcare through AI-driven systems. As we move forward, it is crucial to address privacy and confidentiality concerns to ensure the ethical application of these powerful tools in real-world clinical settings^6^.

**Table 1.**
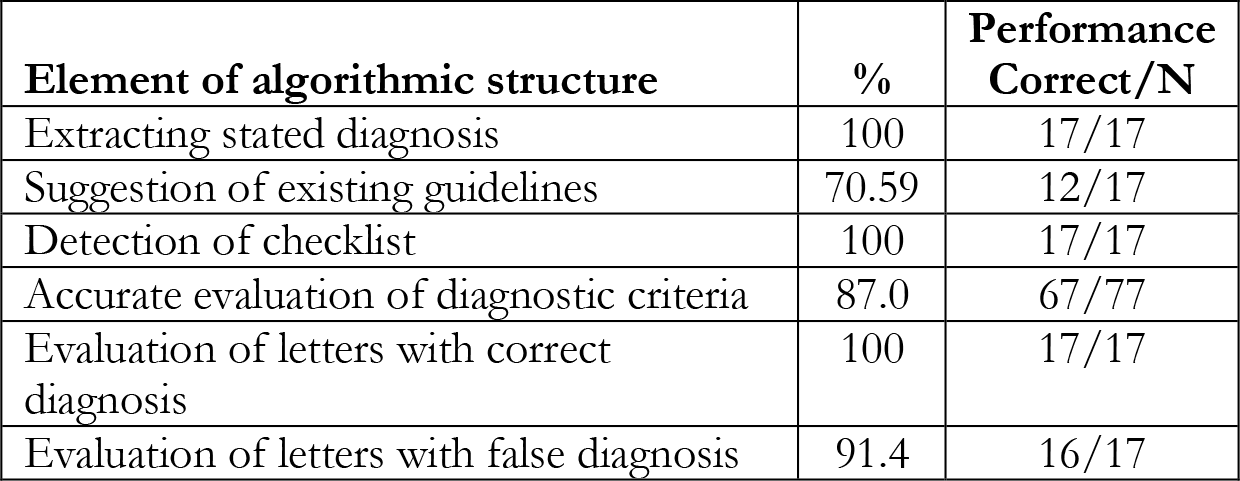
Performance of MedCheckLLM.

## Data Availability

All data produced in the present study are available upon reasonable request to the authors.

**Supplement 1**

Chat Prompts Used in the Study

**Supplement 2**

Data Sharing Statement

