## Supplement 1 for "Large Language Model-Driven Evaluation of Medical Records Using MedCheckLLM"

**Supplement 1**
Prompts Used in the Study

model="**gpt-4-0613**"

**Create Example Medical Report**
{"role": "user", "content": "Please write an example medical report for a person with the diagnosis " + diagnosis + ".\n" }

**Extract Diagnosis**
{"role": "user", "content": "You are supposed to extract a diagnosis of a medical report. Of the choices, return the approriate headache type key for the diagnosis in the following medical report:\n\nKeys:" + KEYS +"\nMedical Report:\n" + report + "\n\n Please return ONLY the key, so that I can access the dictionary directly., e.g.:\n14.1 Headache not elsewhere classified"}

**Select Guideline**
{"role": "user", "content": '''Please examine the following medical report and provide:
- The leading symptom
- potential syndrome
- The diagnosis
- The name of a relevant, established medical guideline for the leading symptom.

\n Do not return actual medical advice but the name of a relevant medical guideline.

\nMedical Report:\n''' + report + '''\n\n'''}

**Turn into Checklist**
{"role": "user", "content": f'''
I will provide you with part of a medical guideline. If it is in a format that could be used as a checklist, return the word 'CHECKLIST'. Only return the word 'CHECKLIST', not the actual checklist itself.
If it is a continuous, long text with complete sentences, try to extract a checklist from the guideline and return your checklist.
\n
Guideline:\n
{guideline_out}
'''}

**Evaluate Letter**
{"role": "user", "content": """
I will provide you with a checklist guideline and a medical report.
Please assess the doctor's letter based on the checklist and consider the following:
Checklist items are usually numbered A,B,C,D,E ...
Understand what is meant by for example: Any headache fulfilling criterion B and C.
Then assess the letter and

return your results:

- For each checklist item separately:

- Whether the checklist item was thoroughly addressed in the doctor's letter (0-5: not covered at all (0) - comprehensively covered (5)).
- Comment.
- Additional comments. '''}

**Correct Diagnosis**

```
{"role": "user", "content": """
    I will provide you with a medical report.
    Please assess whether the doctor's letter identified the correct diagnosis.
     Return the following results:
       - The diagnosis stated in the doctor's letter
       - The diagnosis that you believe the patient actually has
       - whether the stated and actual diagnosis are the same (yes/no)
       - Additional comments.

  """},
  {"role": "user", "content": "Letter:\n" + letter}
```
