## Supplement 2 for "Large Language Model-Driven Evaluation of Medical Records Using MedCheckLLM"

### Data availability statement

No patient data was used in this study. The prompts used in this study are deposited in Supplement 1. Software code used for inquiring the Open AI API and medical reports will be available on GitHub: https://github.com/venkataramani-lab/.
